# Assessment of Digital Payment for Agents in Mass Chemoprevention Campaigns: The Karangue Fay Project in Senegal

**DOI:** 10.1101/2025.03.13.25322936

**Authors:** Jean Augustin Diegane Tine, Amadou Yeri Camara, Aminata Diaw, Meissa Seck, Saliou Séne, Fatoumata Zahra Mohamed Mboup, Amadou Ibra Diallo, Fatoumata Bintou Diongue, Mouhamadou Faly BA, Ibrahima Ndiaye, Souleymane Ndiaye, Adama Faye

## Abstract

**Introduction:** The payment of healthcare agents is a crucial aspect of organizing mass health campaigns. This study aims to examine the effects of digital payments in seasonal malaria chemoprevention (SMC) campaigns in Senegal.

**Methods:** This quasi-experimental study used a three-arm before-after/here-elsewhere design, including (i) a mandatory digital payment zone in Kounkane (Kolda region), (ii) a voluntary digital payment zone in Koussanar (Tambacounda region), and (iii) a control zone without digital payment in Bantaco (Kedougou region). A mixed-method approach combining both qualitative and quantitative data collection was employed from March to June 2023. Ethical approval was obtained from the Senegalese National Ethics Committee for Health Research (CNERS) before field activities began (approval number: 0209 MSAS/CNERS/SP).

**Results:** A total of 299 agents participated in the study, with 181 surveyed before the intervention and 118 after. Participants were distributed as follows: Kounkane (Kolda) 48.8%, Koussanar (Tambacounda) 35.8%, and Bantaco (Kedougou) 15.4%. The key stakeholders included community health workers (90.9%) and supervisors (5.4%) responsible for receiving payments. Among them, 33.5% were single, while 65.2% were married. Male agents constituted 50.8% of the sample. The median age of participants was 32 years, with a median tenure of 3 years. All agents had at least one mobile money account. Most SMC agents used Wave and Orange Money accounts (69.89%), with 90% using Orange Money and 96% using Wave. The main criteria for choosing a reliable money transfer operator included security (95%), ease of use (90.3%), and cash availability (79.2%). Users perceived Karangue Fay as a more secure financial transaction system (82.45%), a reliable process (83.10%), enabling faster transactions (78.20%), more transparent (91.25%), and confidential (95.20%). Compared to cash payments, digital payments were preferred for their transaction security (p<0.001), speed, and ease of use (p<0.001). Qualitative interviews highlighted that digital payments were traceable, fast, and secure; however, reliability depended on stable internet coverage.

**Conclusion:** SMC agents demonstrated strong acceptability of digital payments, contributing to improved campaign coverage across different implementation phases. Additionally, digital payments enhanced community health workers’ engagement and optimized their performance.

## 1. Introduction

The financial autonomy of healthcare providers affects the quality of service delivery and the healthcare system’s ability to meet users’ needs. The more decision-making rights providers have in various areas, the more flexibility they have to respond to purchasing and payment incentives, making these incentives more effective [1]. However, provider autonomy must be accompanied by management capacity, access to information, and accountability [2]. The remuneration of agents involved in mass health campaigns is part of this delicate balance [3]. National or regional mass health campaigns organized by the Senegalese Ministry of Health and Social Action (MSAS) employ a significant number of volunteer community members, referred to as community health workers, who receive financial compensation in the form of allowances. These campaigns frequently rely on cash payments to compensate providers. Large sums of money are transported across different regions, exposing the process to significant security and logistical challenges. Since the early 2010s, many international stakeholders have praised the potential benefits of using mobile phones for healthcare in Africa and have launched various mHealth projects across the continent [4,5].

This study aims to address the limitations of traditional cash payment systems in mass campaigns by introducing a digital payment system in a rural sub-Saharan setting. The Seasonal Malaria Chemoprevention (SMC) campaign was used as a pilot intervention in three regions of southern Senegal (Kolda, Tambacounda, and Kedougou). SMC is an effective public health intervention for reducing the burden of malaria in high-risk regions, particularly for children who are more vulnerable to severe *Plasmodium falciparum* infections [6]. However, it is crucial that SMC is implemented in accordance with World Health Organization (WHO) guidelines and national protocols to prevent drug resistance and adverse effects. To be effective in reducing malaria transmission, it should be integrated with other malaria control interventions, such as the use of insecticide-treated mosquito nets, early diagnosis and treatment of malaria, indoor residual insecticide spraying, and environmental sanitation initiatives [7].

The overall objective of this research is to assess the effects of digital payments in seasonal malaria chemoprevention campaigns in Senegal. Specifically, the study aims to: i) Map payment systems and financial transaction circuits in the study areas; ii) Analyze the perceptions and behaviors of healthcare actors regarding digital payments in health interventions such as SMC; iii) Evaluate the experimental digital payment system “Karangue Fay” used during the SMC campaign in Senegal; iv) Assess the effects of the digital payment strategy on the implementation of the 2023 SMC campaign in Senegal.

## 2. Methods

### 2.1. Study area

Malaria is an endemic disease in Senegal, exposing the entire population to its risks. In 2020, the country accounted for 0.7% of global malaria-related deaths [8]. Between 2017 and 2020, recorded malaria cases decreased by 4.4%, from 52 to 50 cases per 1,000 people at risk. However, during the same period, malaria-related mortality slightly increased by 1.8%, rising from 0.24 to 0.245 deaths per 1,000 people at risk [9].

To combat this public health issue, Senegal’s National Malaria Control Program (PNLP) has implemented various interventions, including the distribution of long-lasting insecticide-treated nets (LLINs), community-based malaria management (PECADOM), and school-based initiatives such as PECA-École and PECA-Daara in traditional and Quranic schools. Another key intervention is Seasonal Malaria Chemoprevention (SMC). Recommended by the World Health Organization (WHO) in 2012, SMC targets children up to the age of ten. Senegal has been a major research site for SMC, initially targeting children under five and later expanding to those under ten.

SMC campaigns are implemented in sixteen districts across the five most affected regions (*Kedougou, Kolda, Tambacounda, Kaolack, and Diourbel*). These interventions cover all districts in Kolda, Kedougou, and Tambacounda, as well as the districts of Touba, Diourbel, and Kaolack. In 2021, monthly treatment rounds were conducted over four months in Kedougou and three months in other target regions. Due to the length of the transmission season in these areas, an additional monthly round was introduced in 2022 to improve coverage during the transmission period [10].

In Senegal, the mobile telecommunications sector generated 428 billion CFA francs in 2020, down from 467 billion CFA francs in 2019, marking a decline of 8.39%. This market was also affected by the COVID-19 pandemic. Despite this, the number of mobile phone lines continued to grow in 2020, reaching 19,078,948 lines, a 6.7% increase compared to 2019 [11]. According to the University of Sherbrooke, internet usage in Senegal grew from 1% in 2002 to 42.6% in 2020 [12]. Additionally, World Bank data indicates that Senegal has one of the highest smartphone adoption rates in West Africa (35.6%), compared to a regional average of 28%. Mobile payment adoption in businesses varies by company size, with 2% in small enterprises, 2.5% in medium enterprises, and 2.8% in large enterprises [13].

The USAID Global Health Finance and Governance (HFG) Project promotes mobile money usage to strengthen health systems [14]. As part of this study, three districts in southern Senegal were selected [15]: Kounkane (Kolda), Koussanar (Tambacounda), Bantaco (Kedougou).

### 2.2. Study Design and data collection

This is a quasi-experimental study using a before-after/here-elsewhere three-arm design: (i) the first intervention arm implemented mandatory digital payments in Kounkane, (ii) the second intervention arm allowed voluntary digital payments in Koussanar and (iii) the control arm used no digital payment in Bantaco.

### 2.3. Data Collection and Analysis

A mixed-method approach combining both qualitative and quantitative methodologies was used for data collection.

Quantitative data were collected from all agents involved in the Seasonal Malaria Chemoprevention (SMC) campaign in the study areas before and after the deployment of the Karangue Fay digital payment platform. Categorical variables were described using absolute and relative frequencies. Continuous variables were described using Mean and standard deviation for parametric distributions and Median and interquartile range (IQR) for non-parametric distributions. Statistical tests and multivariate analyses were conducted in accordance with the required application conditions. The RStudio software [16] was used for data analysis.

Qualitative data collection employed three methods: Semi-structured individual interviews, Focus group discussions and Informal interviews.

The qualitative data collection process was iterative, involving: An initial field visit before the intervention, A second field visit after the intervention and Ongoing informal interviews throughout the analysis, using an empirical-inductive approach. Qualitative data collected in the field were processed using Nvivo software [17].

### 2.4. Ethical Considerations

Ethical approval was obtained from the Senegal National Ethics Committee for Health Research (CNERS) [18] before field activities began (approval number 0209 MSAS/CNERS/SP). Additionally, authorization was obtained from the Ministry of Health and Social Action through the National Malaria Control Program (PNLP) before the study commenced. The medical authorities of the Kolda, Tambacounda, and Kedougou regions, as well as district health officials, were briefed on the project and actively involved in its implementation. Participation in this study was voluntary. Informed consent was obtained from all participants. An information sheet was provided to each participant before signing the consent form.

### 2.5. Description of the Karangue Fay Project

The Karangue Fay project was submitted under the Digital Health Payment Initiative and Research (DHPI-R) project [19], funded by Bill and Melinda Gates and aims to generate evidence on digital payment for rural health workers and establish its impact on the delivery of health campaigns in sub-Saharan Africa.

As part of this project, the Karangue Fay (KF) platform was developed through a participatory and iterative approach involving stakeholders from the Ministry of Health and Social Action (MSAS), the University Cheikh Anta Diop (UCAD), and community actors between March and June 2023.

The platform is a web application, developed with local expertise, capable of enabling money transfers through three payment methods: Wave [20], Orange Money [21], and Free Money [22]. It also allows transaction tracking and payment status generation while ensuring multi-level security, which will be described later. Access to the platform was provided through the URL www.karanguefay.sn, after pre-registration of users by the system developer. Authorization levels varied among users based on their roles and responsibilities within the healthcare system. Regional Health Directors (DRS) and Regional Managers (GR) were responsible for overseeing district-level operations, while District Chief Medical Officers (MCD) supervised health post payments remotely from their offices. District Managers (GD) managed the platform by updating the list of healthcare agents eligible for payment. At the health post level, Head Nurses (ICP) ensured the accuracy of payment requests by verifying names and amounts before approval. The Karangue Fay platform administrator had full access rights, allowing them to monitor transactions and cancel duplicate payments when necessary. Access to the platform was secured through a two-step authentication process: (i) Email login and password entry, (ii) A confirmation code sent via SMS.

The beneficiary’s information including first name, last name, national identification number, phone number, and preferred mobile money operator, was recorded on the platform.

Afterwards, the transfer amount was assigned to each beneficiary. A payment confirmation message was sent via text message, stating: “You have received XX,000 FCFA via Orange Money (or Wave or Free Money), sent by Karangue Fay”.

A payment report was generated in PDF format, including the logos of partner organizations and validated activity titles. This report was then sent via email to the various managers and was also available for direct download from the platform.

## 3. Results

The total number of agents who participated in the study was 299, with 181 in the pre-intervention phase and 118 in the post-intervention phase. This difference is explained by the exclusion of certain SMC agents following the results obtained during the preliminary training before their field enrollment.

The participants were residents of Kounkane at 48.8%, then of Koussanar at 35.8% and of Bantako 15.4%. The key actors involved were predominantly community health workers (90.9%), followed by supervisors (5.4%). Regarding marital status, 65.2% of the participants were married, while 33.5% were single. Men constituted a slight majority, representing 50.8% of the participants. The median age of participants was 32 years, and their median tenure in their roles was 3 years.

### 3.1. Mapping of Payment Systems and Financial Transaction Circuits

The mapping of digital platforms allowed us to understand the feasibility of digital payments in the study area. Mobile phones were the most commonly used tool among SMC agents, with 99% of healthcare agents owning a functional mobile phone. For the Karangue Fay project, having a mobile phone was essential for payment verification. A total of 98% of agents were already subscribed to a mobile network operator that enabled digital money transactions. Wave was the preferred payment method across all intervention areas, and cash withdrawal was the primary transaction conducted. Bank account coverage was low across the three research sites. The table I presents the availability of digital payment options.

**Table I:**
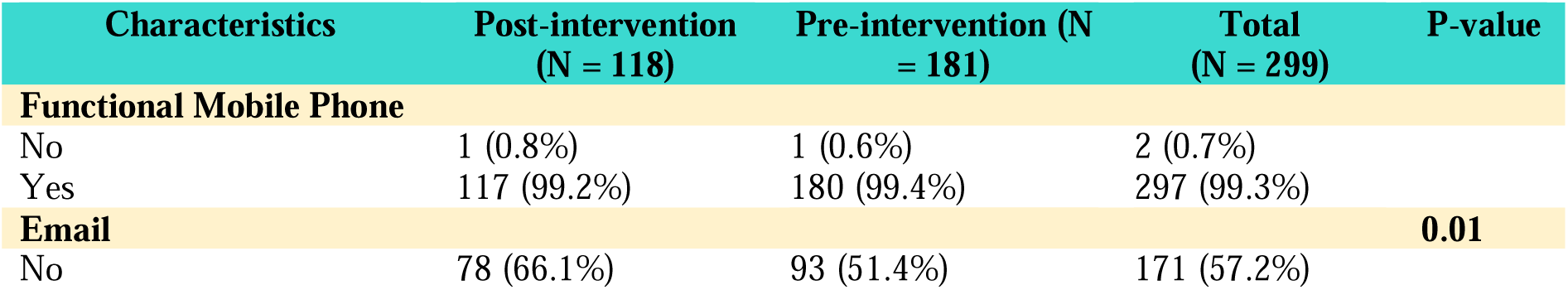
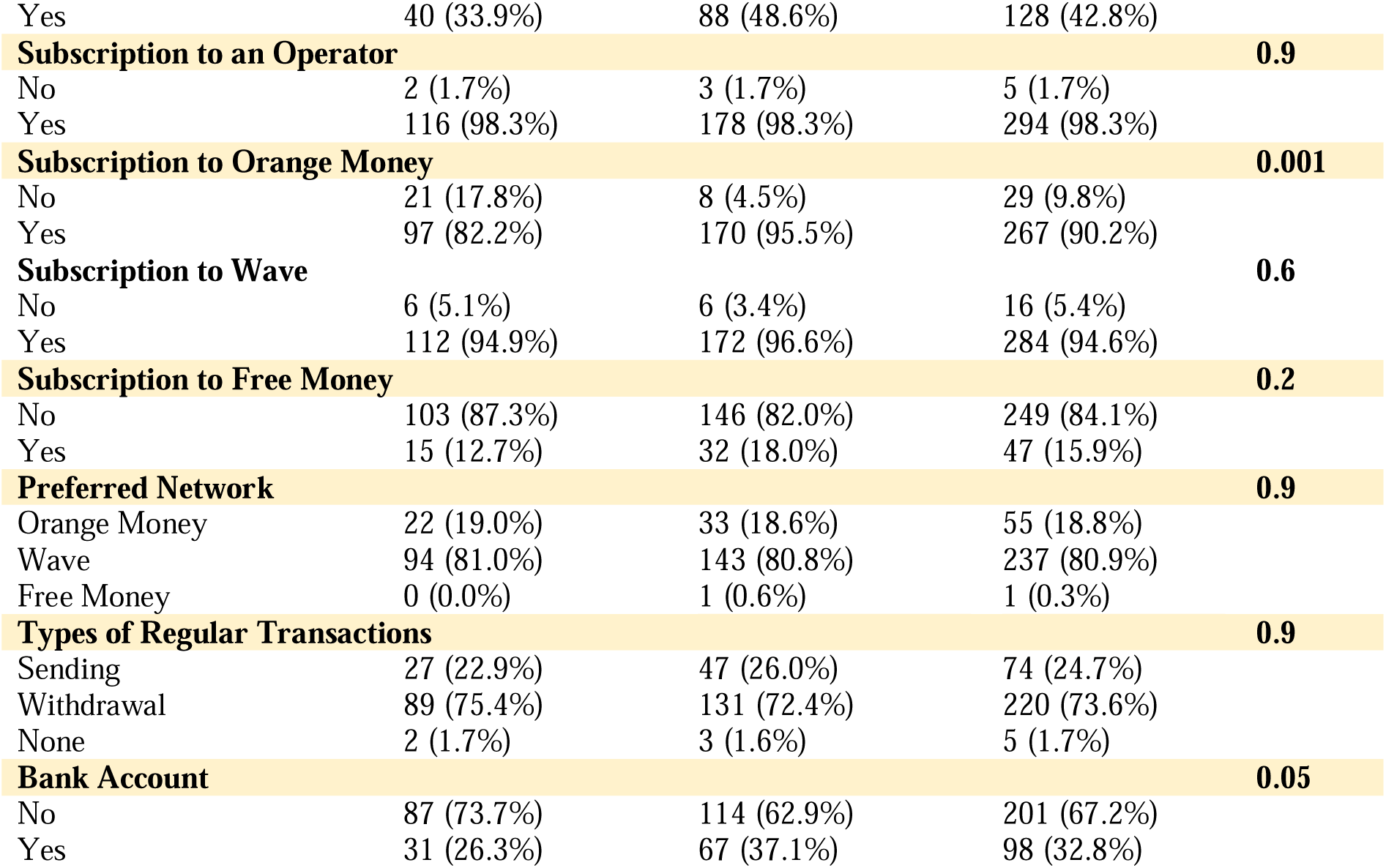
Availability of Financial Exchange Means and Equipment.

### 3.2. Perceptions of the Functioning of Different Payment Methods

In the study area, several payment methods were identified: cash payment, bank transfer, check payment, and digital payment. Among these options, cash payment remains the most traditional method. Figure 1 illustrates the evolution of beneficiaries’ perceptions regarding these different payment methods before and after the intervention. The results highlight that cash payments are perceived as less confidential, with longer waiting times. Payments made via bank transfer or check are also considered less efficient due to the time constraints imposed by banks’ opening and closing hours.

**Figure 1:**
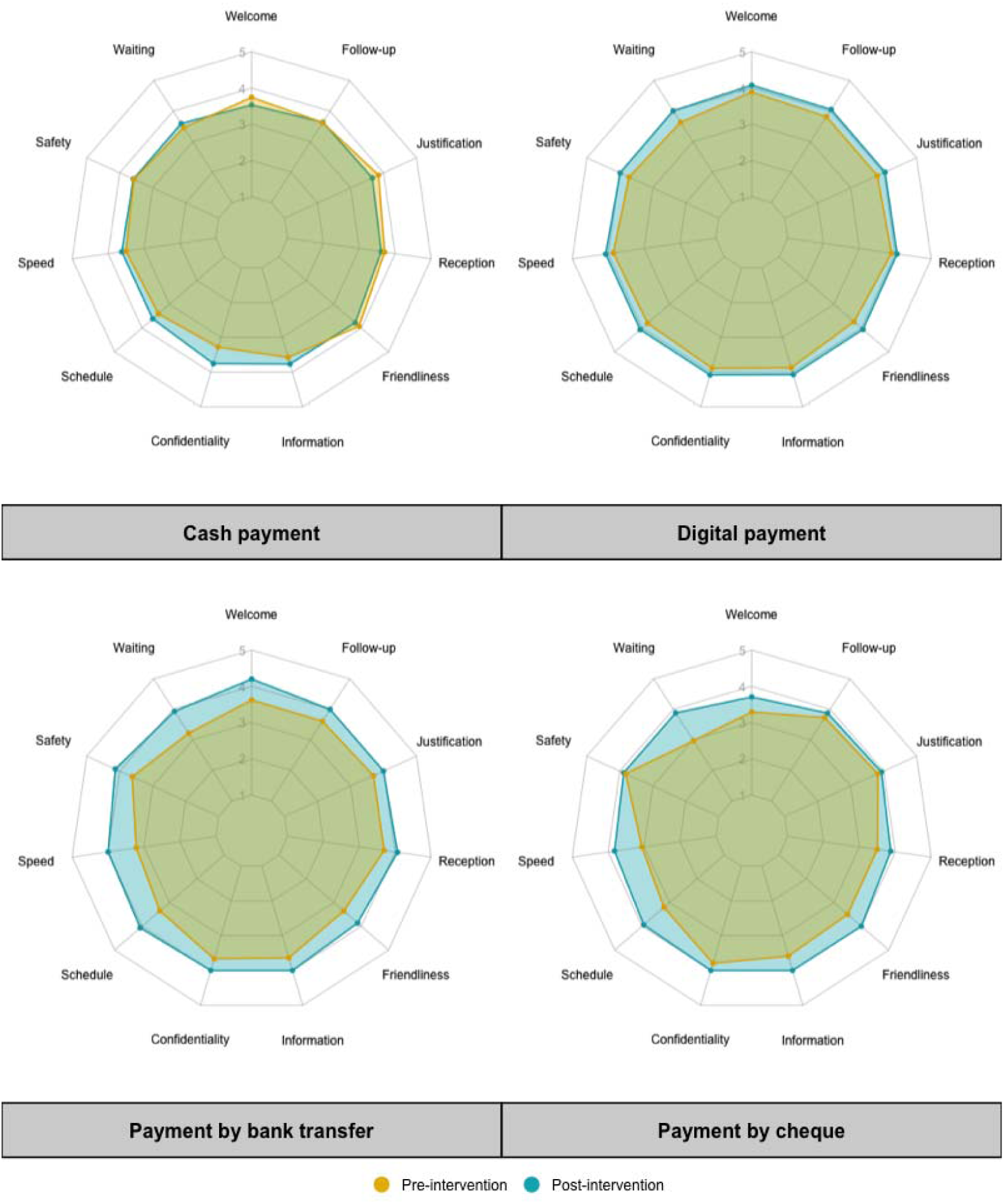
Synoptic Representation of the Appreciation Levels of the Qualities of Different Payment Methods Throughout the Project.

Conversely, digital payment received the highest satisfaction scores across the eleven evaluated dimensions, both before and after the intervention. Respondents particularly emphasized speed, convenience (notably the user-friendliness of the process), and traceability (ease of justification) as the main reasons for their satisfaction with this payment method.

Qualitative interviews provided deeper insights into perceptions of different payment methods. Although cash payment is widely used and well understood by managers, users acknowledge that it has significant drawbacks, such as increased vulnerability to theft and money loss. However, this payment method avoids the risks associated with online fraud, which is a notable advantage. Bank transfers and check payments are generally perceived as more secure options. Nevertheless, these payment methods involve operational constraints, including extended waiting times at banks. Additionally, difficulties may arise due to administrative requirements, such as signatures and identification documents. A nurse from Kounkane shared: *“I have often encountered errors in the spelling of my first name, which caused difficulties in receiving my money at the bank “.* Other participants also reported input errors that led to similar complications in the banking process. Finally, digital payments, while equipped with advanced security protocols, require a reliable transfer network to ensure their efficiency and accessibility.

### 3.3. Assessment of the Costs of Different Payment Methods

Different payment methods incur direct or indirect additional costs. Cash payments generate extra transportation expenses and potential income loss due to waiting times. Bank transactions lead to account maintenance fees and the purchase of transaction accessories (such as bank cards and checkbooks). For digital payments, costs are associated with transactions, but they require the user to have a functional mobile phone and an active phone subscription. Some digital payment services charge fees, such as transaction or currency conversion fees. These costs can accumulate, particularly for international payments. The cost assessment by payment method and site (Figure 2) indicated a higher acceptance of digital payment costs compared to other payment methods. Digital payments reduce the expenses incurred when retrieving cash and the time lost due to waiting.

**Figure 2:**
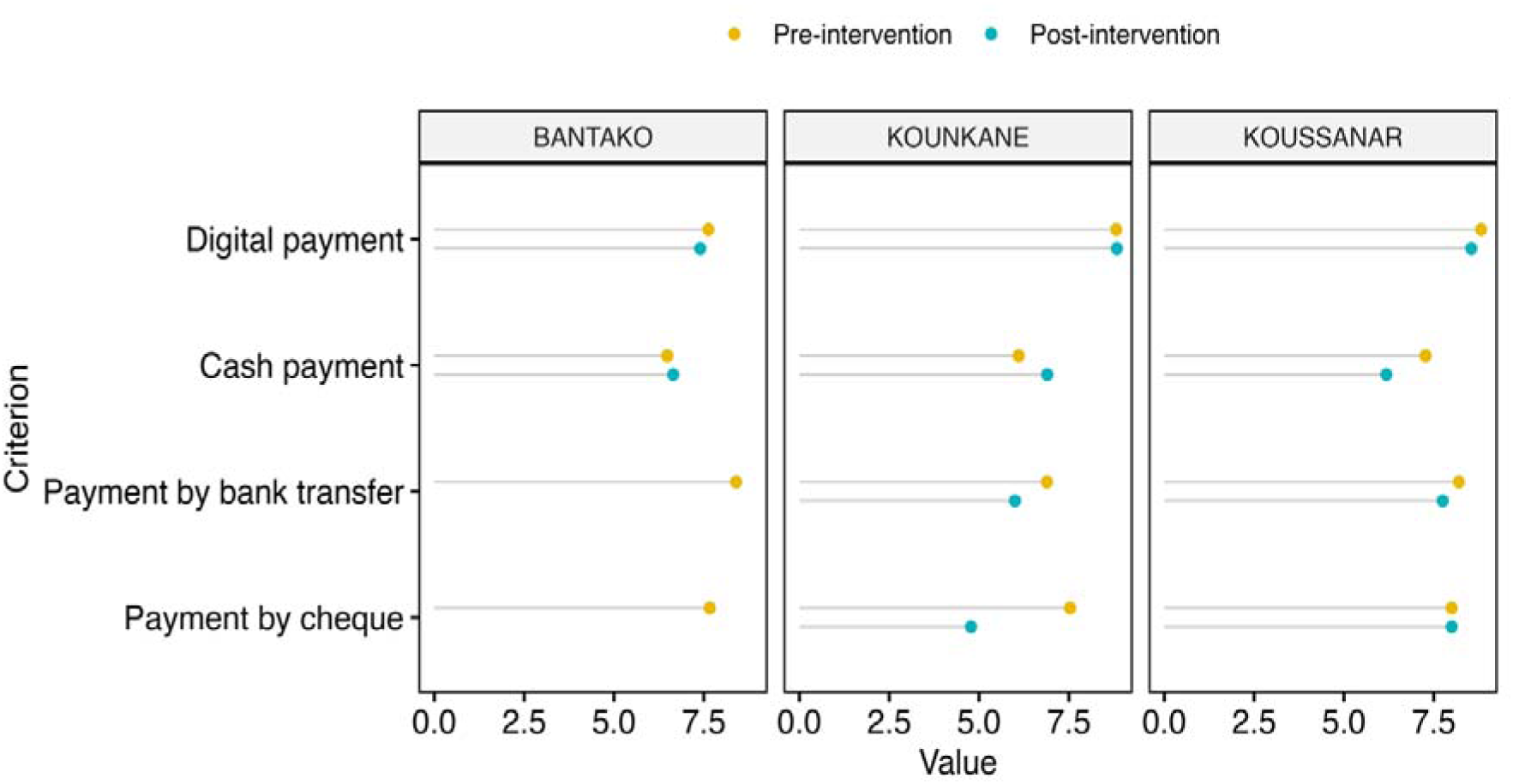
Preferential Assessment of Payment Methods Based on Costs (Direct and Indirect) on a Scale from 0 to 10.

### 3.4. Assessment of User Satisfaction with the Karangue Fay Platform

The project assessed preferences for digital payment across multiple dimensions. Head Nurses (ICP) and community health workers particularly appreciated the time and resource savings offered by this payment method. Since transactions were conducted remotely, the ICPs were able to continue their healthcare activities without interruption, while community health workers did not have to pause their usual tasks to receive their payments. Moreover, beneficiaries did not face difficulties related to finding change, a constraint that often kept them waiting late at health posts. Most respondents reported having waited several days to receive their payments; some even had to travel multiple times before receiving their funds. One community health worker shared: *“Yes, I had to go to the Koussanar health post to receive the payment. I didn’t have a problem because I live 4 kilometers away, and I have a motorcycle, so the distance wasn’t an issue for me. But with cash payments, we often face difficulties because sometimes we travel to the post, and the money isn’t available “*.

These observations confirm that digital payment facilitates better time and financial resource management, meeting beneficiaries’ expectations while improving their efficiency in carrying out their tasks. A community supervisor shared in an individual interview: *“Yes, we saved time. Personally, digital payment was very convenient for me because with cash payments, sometimes I was the one collecting the funds from the district, and at times, I was also responsible for making the payments. As a result, after the campaign, I would spend over three days managing payments. But during this last round, I finished work on Saturday, and on Sunday, I dropped off the materials at the post—at that moment, I received the payment notification “*.

The overall satisfaction assessment regarding the digital payment system implemented by Karangue Fay revealed that 88.9% of users reported being completely satisfied. This satisfaction rate was 87% in Kounkane and 90.5% in Koussanar (see Table 2). Additionally, the overall acceptance of using the platform for future payments was 97.3%, with 97.4% in Kounkane and 97.2% in Koussanar.

**Table II:** Satisfaction Level and Acceptability of Beneficiaries Regarding the Karangue Fay (KF) Digital Payment System.

| Items | Kouankane (N=78) | Koussanar (N=74) | Total (N=152) |
| --- | --- | --- | --- |
| **Satisfaction by Dimension** |  |  |  |
| Reliable | 65 (83.0%) | 62 (83.2%) | 126 (83.1%) |
| Secure | 63 (81.1%) | 62 (83.8%) | 125 (82.4%) |
| Fast | 60 (77.3%) | 59 (79.1%) | 119 (78.2%) |
| Transparent | 70 (90.3%) | 68 (92.2%) | 139 (91.2%) |
| Confidential | 73 (94.0%) | 71 (96.4%) | 145 (95.2%) |
| **Overall Satisfaction** |  |  |  |
| Very Satisfied | 68 (87.2%) | 67 (90.5%) | 135 (88.9%) |
| Satisfied | 4 (5.1%) | 5 (6.8%) | 9 (5.9%) |
| Somewhat Satisfied | 4 (5.1%) | 2 (2.7%) | 6 (3.9%) |
| Not Satisfied | 2 (2.6%) | 0 (0.0%) | 2 (1.3%) |
| Not at All Satisfied | 0 (0.0%) | 0 (0.0%) | 0 (0.0%) |
| **Acceptability of Using the KF Platform for Future Campaigns** |  |  |  |
| Yes | 76 (97.4%) | 72 (97.2%) | 148 (97.3%) |
| No Opinion | 2 (2.6%) | 1 (1.4%) | 3 (2.0%) |
| No | 0 (0.0%) | 1 (1.4%) | 1 (0.7%) |

The findings do not indicate a specific profile associated with the adoption of digital payment within the SMC campaigns. It appears that adherence to this payment method is more influenced by technical and organizational factors rather than individual characteristics.

Despite the predominantly positive feedback collected from various stakeholders, certain limitations were identified at the two sites where payments were processed. The main challenges involved transaction failures and cancellations. In Koussanar, respondents reported six cases of transaction failures. According to the information gathered, these failures were attributed to: two number entry errors, one inactive phone number, and three cases where community health workers had selected a payment operator without having an active account with that operator.

The ICP of Koussanar explained: *“In my opinion, these are minor details, as they were related to form-filling issues. We observed errors in number entry, often due to a lack of familiarity among community health workers, for instance, when they provided incorrect numbers or mistakenly used 77 instead of 76 at the beginning of a number “*.

These findings highlight the critical importance of raising awareness among stakeholders about the digital payment process. Such awareness efforts are essential to ensuring the successful implementation of this payment method during large-scale health campaigns.

### 3.5. Impact of the Digital Payment Strategy on the Implementation of the 2023 SMC Campaign in Senegal

Formally establishing a direct impact of the Karangue Fay project on malaria-related morbidity in Senegal remains challenging. However, performance indicators assessing the coverage of seasonal malaria chemoprevention (SMC) campaigns, as recorded at the health posts where the project was implemented, are presented in Table 3.

**Table III:** Results of coverage rates in SMC 2023 campaign by Zone.

| SMC Performance Indicators | Kounkane | Koussanar | Bantako |
| --- | --- | --- | --- |
| Registered Children / Treated Children | 386/390 | 4202/4353 | 1414/1485 |
| Coverage for DOT-1 | 99% | 96.5% | 95.2% |
| Coverage for DOT-2 | 99% | 96.5% | 95.4% |
| Coverage for DOT-3 | 99% | 96.5% | 95.4% |
| Coverage for All 3 Rounds | 100% | 100% | 99.5% |

*DOT-1: Directly Observed Treatment – First Round*

The data reveal higher SMC coverage rates in Kounkane and Koussanar compared to the control area of Bantako. These results suggest that the benefits of digital payment— particularly its speed, traceability, and the streamlining of administrative procedures—may contribute to increased engagement among community health workers, ultimately enhancing their effectiveness in executing public health campaigns.

## 4. Discussion

Over the past decade, digital technology has gained renewed interest with the widespread adoption of mobile phones and mobile internet across the African continent. Africa now hosts the largest number of mobile development programs, or *mDevelopment*, positioning itself as a *new digital frontier* driven by its mobile revolution [23,24].

The digitalization of payments involves the complete dematerialization of financial transactions, including both revenue collection and expenditures [25]. A digital environment necessitates a strong commitment to providing clients with transparent information about financial products, particularly when such information is accessible only through electronic means, such as mobile technology [26]. This was a key consideration in the Karangue Fay pilot project, whose preliminary objective was to assess the digital environment of the study area and evaluate the feasibility of implementing digital payment systems.

The study zones are rural areas with generally low living standards. However, mobile phone ownership was found to be 99%, with 98% of participants subscribed to a mobile money operator. Moreover, 74% of participants had prior experience with digital payments, particularly for withdrawals. The rapid expansion of mobile telephony is closely linked to the limited penetration of fixed-line telephony in sub-Saharan African countries (3% in Senegal, 1.2% in Côte d’Ivoire, and 0.9% in Burkina Faso) [5]. In 2020, 46.2% of the Senegalese population had internet access, though usage remained concentrated in major urban areas [11,12]. Most users accessed the internet via 3G mobile networks, but limitations in 3G and 4G coverage, as well as the high cost of internet access and mobile communication services, restricted usage to wealthier social classes [5,12].

Digital health technologies have the potential to be cost-effective tools that improve healthcare quality, optimize expenditures, and enhance patient-provider interactions [3,27,28]. In many African countries where bank account penetration is relatively low (33% in our study), mobile banking presents a significant opportunity for digital health. It enables individuals to pay for healthcare expenses via mobile money while also facilitating the remuneration of healthcare workers.

In this project, overall satisfaction with digital payment was 94.8%, with 88.9% of users reporting complete satisfaction. Digital transactions were perceived as less costly than the indirect expenses associated with other payment methods, such as cash or bank transfers. Acceptance of the Karangue Fay digital payment platform was 97.3%, and health workers expressed a strong preference for continuing this payment method in future malaria chemoprevention campaigns.

Since 2021, Senegal has launched the “eSanté “ initiative, a national program for the digitalization of the healthcare system, covering both healthcare professionals and patients over the 2022–2027 period [29]. This program aims to integrate digital technologies and platforms into the healthcare system to enhance service delivery and health governance.

This study found no significant socio-cultural or economic barriers to the adoption of digital payment by healthcare workers. However, structural challenges were identified. Six transaction failures out of 154 transfers were documented, primarily due to errors in beneficiary information (e.g., incorrect phone numbers or selection of an inactive mobile money operator). Additional challenges included internet connectivity issues and the availability of cash at mobile money withdrawal points.

For digital payment systems to be fully effective, seamless financial transaction management is essential. This requires a well-coordinated synergy between internet service providers, mobile money operators, local merchants, and end-users [30].

The adoption of mobile financial services in insurance and healthcare is closely linked to building a sustainable and profitable mobile economy in Africa, where 90% of users purchase mobile credit on a pay-as-you-go basis and frequently switch service providers. In this context, eHealth solutions can serve as loyalty programs, helping service providers retain customers in an increasingly competitive and unstable market [31].

The high acceptability of digital payments among health workers involved in SMC campaigns contributed to improved campaign coverage across multiple implementation rounds. This, in turn, strengthened the engagement of community health workers and enhanced their performance.

The World Health Organization’s (WHO) Global Strategy on Digital Health (2020–2025) states that:

*“Digital health must be an integral part of healthcare priorities, ensuring ethical, safe, reliable, equitable, and sustainable benefits for individuals. It should be developed based on principles of transparency, accessibility, scalability, replicability, interoperability, privacy, security, and confidentiality “* [26]. These are precisely the key attributes and values expected of digital payment systems.

### Study Limitations

This pilot study did not have access to internet service providers to assess the digital divide, which measures difficulties in accessing the internet. Additionally, the impact of digital payment on malaria control could not be objectively determined due to the limited timeframe. However, SMC coverage performance improved in the two implementation zones.

Given this favorable digital environment, we believe that adopting digital payment as the primary payment method could provide health system stakeholders with a more direct, faster, and more secure means of payment. However, to scale up this payment method, further exploration is needed to understand how it can be integrated into a broader range of public health activities.

## 5. Conclusion

In its WHA58.28 resolution on eHealth, the World Health Organization (WHO) urged Member States *“to consider developing a long-term strategic plan for designing and implementing eHealth services across various areas of the health sector […] to promote equitable, affordable, and universal access to their benefits”* [32].

According to the Ministry of Health and Social Action (MSAS), Senegal’s eHealth digitalization program aims to provide users with accurate health information, enhance the quality and safety of care, expand the range of medical specialties available to healthcare workers, and strengthen the connection between patients and healthcare providers [33].

The transition to digital health in Senegal represents a promising opportunity, yet it requires a comprehensive understanding of the challenges associated with this shift. Our study, which examines the use of digital payments in community health activities, highlights several key insights. It reveals that health providers overwhelmingly own functional mobile phones and are predominantly subscribed to mobile money services, particularly Wave and Orange Money.

Digital payments are perceived as fast, secure, traceable, and convenient, although their efficiency remains contingent upon network coverage and the availability of withdrawal points. The Karangue Fay platform, considered reliable, transparent, and confidential, has enhanced the acceptability of digital payments and contributed to improving the performance of community health activities, particularly in the project implementation areas.

These findings underscore the potential of digital tools to enhance the effectiveness of health interventions in Senegal.

## Data Availability

We confirm that all data underlying the conclusions of this study are fully available without restriction, in accordance with PLOS policies.

## Notes

### Competing Interest Statement

The authors have declared no competing interest.

## Références

1. Cashin C. Assessing Health Provider Payment Systems: A Practical Guide for Countries Working Toward Universal Health Coverage [Internet]. Joint Learning Network for Universal Health Coverage; 2015. Available at: https://www.jointlearningnetwork.org/wp-content/uploads/2019/11/AT_Workbook_completePDF.pdf

2. Trisnantoro L, Hendrartini J, Susilowati T, Miranti PAD, Aristianti V. A critical analysis of selected healthcare purchasing mechanisms in Indonesia [Internet]. World Health Organization; 2016 [cited Dec 19, 2024], p. 96-151. *(Strategic Purchasing in China, Indonesia, and the Philippines).* Available at: https://www.jstor.org/stable/resrep28440.5

3. Al Dahdah M. Digitalization of Healthcare in the Global South: When Digital Firms Decide on Access to Care. Mouvements. 2019;98(2):120–32.

4. Mechael P, Batavia H, Kaonga N, Searle A, Kwan A, Goldberger L, et al.Barriers and Gaps Affecting mHealth in Low and Middle-Income Countries: Policy White Paper [Internet]. mHealth Alliance; 2010. Available at: https://www.mobileactive.org/pdfs/mHealth_Barriers_White_Paper.pdf

5. Weil O, Tikkanen M, Kouanda S, Absolu A. French Development Agency. 2013 [cited Dec 19, 2024]. The Use of Information and Communication Technologies (ICTs) in Maternal and Child Health in Sub-Saharan Africa. Available at: https://www.afd.fr/fr/ressources/lutilisation-des-nouvelles-technologies-de-linformation-et-des-communications-tic-dans-le-domaine-de-la-sante-maternelle-et-infantile-en-afrique-subsaharienne

6. Diop S, Kaly J, Lawson D, Diop M, Diop B. *Knowledge, Attitudes,* and Practices of Mothers or Caregivers Regarding Seasonal Malaria Chemoprevention. Médecine Mal Infect. June 1, 2017;47(4, Supplement):S97.

7. World Health Organization. WHO General Policy Recommendation: Seasonal Malaria Chemoprevention for Controlling Plasmodium falciparum Malaria in Areas of High Seasonal Transmission in the Sahel Subregion of Africa [Internet]. Geneva: World Health Organization; 2012. Available at: https://iris.who.int/handle/10665/337982

8. World Health Organization. World Malaria Report 2021 [Internet]. [cited Dec 20, 2024]. Available at: https://www.who.int/teams/global-malaria-programme/reports/world-malaria-report-2021

9. National Malaria Control Program (PNLP). National Strategic Plan for Malaria Control in Senegal 2016-2020 [Internet]. MSAS; 2020. Available at: https://malariaportal.org/sites/default/files/2023-11/SNG-501.1_%20Plan%20Strategique%20National%20PNLP%202016-2020.pdf

10. U.S. President’s Malaria Initiative - Senegal Malaria. *Operational Plan FY 2022.* USAID [Internet]. 2022. Available at: https://d1u4sg1s9ptc4z.cloudfront.net/uploads/2022/01/FY-2022-Senegal-MOP.pdf

11. ARTP Senegal. *Reports on the Electronic Communications Market.* [cited Nov 27, 2023]. Available at: https://artp.sn/publications/rapports-sur-le-marche-des-communications-electroniques

12. World Perspective. Major Global Trends Since 1945 [Internet]. [cited Nov 27, 2023]. Available at: https://perspective.usherbrooke.ca/bilan/servlet/BMTendanceStatPays

13. World Bank. Senegal - Overview. [cited Oct 22, 2023]. Available at: https://www.banquemondiale.org/fr/country/senegal/overview

14. USAID. Health Finance & Governance Project [Internet]. [cited Nov 27, 2023]. Available at: https://www.hfgproject.org/

15. National Agency for Statistics and Demography (ANSD) of Senegal [Internet]. [cited Dec 20, 2024]. Available at: https://www.ansd.sn/regions

16. Posit [Internet]. [cited Dec 20, 2024]. Available at: https://www.posit.co/

17. Jackson K, Bazeley P. Qualitative Data Analysis with NVivo. 3rd ed. Los Angeles, London, New Delhi, Singapore, Washington, DC, Melbourne: Sage; 2019. 349 p.

18. MSAS. National Health Research Ethics Committee [Internet]. [cited Oct 22, 2023]. Available at: https://www.cners.sn/

19. DHPIR. What is the Digital Health Payment Initiatives & Research in Africa (DHPI-R)? [Internet]. [cited Nov 27, 2023]. Available at: https://dhpir.mak.ac.ug/about-us

20. Wave Mobile. Wave [Internet]. [cited Dec 23, 2024]. Available at: https://www.wave.com/fr/

21. Orange Money. *Orange Money Europe Money Transfer - Send Money to Senegal* [Internet]. 2024 [cited Dec 23, 2024]. Available at: https://orangemoney.fr/transferer-de-l-argent-au-senegal/

22. Free Money. *Free Money – Free* [Internet]. 2024 [cited Dec 23, 2024]. Available at: https://www.free.sn/free-money/

23. Mobile Phones: The New Talking Drums of Everyday Africa [Internet]. Langaa RPCIG; 2009 [cited Dec 24, 2024]. Available at: https://www.jstor.org/stable/j.ctvk3gmgv

24. Cheneau-Loquay A. Proparco Review. 2009 [cited Dec 24, 2024]. *Mobile Telephony in Developing Countries: What Are the Economic and Social Impacts?* Available at: https://www.proparco.fr/fr/ressources/la-telephonie-mobile-dans-les-pays-en-developpement-quels-impacts-economiques-et-sociaux

25. Better Than Cash Alliance. *Eight Principles for Digital Transformation of Public Health - PAHO/WHO | Pan American Health Organization* [Internet]. [cited Nov 26, 2023]. Available at: https://www.paho.org/en/is4h-information-systems-health/8-principles-digital-transformation-public-health

26. WHO. *Global Strategy on Digital Health 2020-2025 [Internet]. [cited Nov 27, 2023]*. Available at: https://www.who.int/fr/publications-detail/9789240020924

27. Ridde V. Access to Healthcare in West Africa [Internet]. Montreal: PUM; 2012 [cited Dec 24, 2024]. *(PUM)*. Available at: https://pum.umontreal.ca/catalogue/acces-aux-soins-de-sante-en-afrique-de-louest-l

28. Sen G, Östlin P, eds. Gender Equity in Health: The Shifting Frontiers of Evidence and Action. New York: Routledge; 2009. 340 p.

29. Dia IK. *UNDP.* [cited Dec 24, 2024]. *e-Health: The Digital Revolution for a Connected Healthcare System.* Available at: https://www.undp.org/fr/senegal/histoires/e-sante-la-revolution-numerique-pour-une-sante-connectee

30. Paillès JC. Electronic Payment Systems on the Internet. Cahiers Numériques. 2003;4(1):45–69.

31. GSMA. *The Mobile Economy Sub-Saharan Africa* 2023 [Internet]. *The Mobile Economy.* [cited Dec 24, 2024]. Available at: https://www.gsma.com/solutions-and-impact/connectivity-for-good/mobile-economy/sub-saharan-africa/

32. WHO. *Seventy-First World Health Assembly* [Internet]. [cited Nov 27, 2023]. Available at: https://apps.who.int/gb/ebwha/pdf_files/WHA71-REC1/A71_2018_REC1-fr.pdf

33. CSSDOS. *Digital Health Strategic Plan 2018-2023 - Senegal* [Internet]. [cited Nov 27, 2023]. Available at: https://www.sante.gouv.sn/Pr%C3%A9sentation/plan-strategique-sante-digitale-2018-2023

